# Mouth-related functional problems and vulnerability among community-dwelling older Canadians: evidence from the Canadian Health Survey on Seniors

**DOI:** 10.64898/2026.07.20.26358509

**Authors:** Vishnu Sreevalsan Menon, Marziyeh Shafizadeh, Anil Menon

## Abstract

**Introduction:** Community-dwelling seniors in Canada face many challenges, including declining oral function, that may substantiate vulnerabilities associated with aging in place like social isolation, impaired mobility, and greater care needs. This study analyzes the effects of functional mouth problems, namely discomfort while eating and avoiding foods while eating due to mouth problems, and additionally poor/fair perceived oral health, on the vulnerability outcomes of loneliness, fear of falling, and receipt of informal home care associated with aging in place among Canadian Seniors.

**Methods:** Secondary analysis of the Canadian Health Survey on Seniors (CHSS), which included information on oral health, socioeconomic status, and daily habits among over 42,000 respondents, allowed assembly of a 32,756-respondent sample set and survey-weighted population estimate post-exclusion. Primary Analysis, using descriptive and regression methods, examined the effects of experiencing discomfort while eating, whilst secondary analysis examined avoidance of foods and perceived oral health values on senior vulnerability outcomes.

**Results:** Both study analyses revealed that all variables associated with oral function exposure were linked to high loneliness and fear of falling, and to receiving informal home care, with the exception of a weaker association between receipt of informal home care and perceived oral health.

**Discussion:** Other studies of oral function and its effects on the vulnerability of the senior population reported findings comparable to this study, indicating that oral functional problems are associated with generally higher vulnerability outcomes.

**Conclusion:** Discomfort while eating and avoiding foods may be potential indicators of vulnerability associated with aging in place among Canadian seniors.

## Background

Canada’s population is undergoing a pronounced aging process, elevating the health, autonomy, and well-being of older adults as a growing public health priority. Current demographic data indicates a significant segment of the population is already aged 65 or older, with a further considerable portion approaching this life stage.^1^ This trend is projected to persist. According to estimates from the Canadian Institute for Health Information, the country’s senior population is anticipated to increase by 68% from 2017 to 2037, ultimately encompassing about 10.4 million individuals.^2^

### Vulnerability of Canadian Seniors and Aging in Place

The expansion of the senior demographic underscores the need to identify factors associated with vulnerability among those aging in place. In the Canadian context, the vulnerability of older adults correlates with social isolation and functional limitations affecting mobility.^3–5^ Loneliness and social isolation are relevant to aging in place as living in one’s community requires both physical independence and sustained social interaction and involvement. An estimated 30 percent of Canadian seniors are considered at risk for social isolation, with 19 to 24 percent reporting actual experiences of isolation or loneliness.^3^ Fear of falling is indicative of a lack of mobility, which is associated with increased difficulties and lower quality of life among those who are community-dwelling.^5^ Additional indicators, namely the receipt of informal home care, may also signify vulnerability, as it often reflects heightened requirements for health or functional support among community-dwelling older adults.^4^

### Oral Health and Mouth-Related Functional Problems

Oral health and proper oral function constitute a significant dimension of health in senior populations, and their absence is associated with adverse outcomes.^6,7^ Existing literature identifies several obstacles to oral care for this demographic, encompassing restricted access to dental services, unfavourable attitudes toward oral health maintenance, challenges related to caregivers, and functional impairments that can hinder both daily oral hygiene routines and the pursuit of professional dental treatment.^6,8–11^

Connections between oral health and broader vulnerability may also operate through functional mechanisms. Research on frailty indicates that oral issues, including masticatory difficulties, salivary dysfunction, and other orofacial disorders, correlate with higher frailty levels in older populations.^12^ Furthermore, deficient oral health status is linked to adverse outcomes such as diminished quality of life, increased disability, and higher mortality risk in seniors.^7,13^ This evidence suggests that oral health likely reflects more than a localized dental issue; it may also indicate broader vulnerability in advanced age.

Mouth-related functional impairments warrant particular attention, as they disrupt essential daily activities such as eating and nutritional intake. Dietary factors and nutritional risks have similarly been associated with mortality and diminished life expectancy in Canada.^14,15^

Dysphagia and other oral or swallowing impairments are prevalent among older adults and can hinder comfortable eating and the maintenance of sufficient nutrition.^16,17^ Consequently, mouth-related issues, including discomfort during eating, food avoidance due to oral problems, and poorer overall oral health, may serve as functional indicators linking oral health to broader vulnerability.

### Knowledge Gap and Canadian Context

Research has confirmed associations between oral health, nutrition, frailty, disability, and mortality,^7,11–13^, but the relationship between mouth-related functional problems and specific vulnerability outcomes in a Canadian senior population remains less understood. This gap warrants attention because these outcomes reflect distinct aspects of vulnerability associated with aging in place in Canada.^18^ Loneliness indicates psychosocial vulnerability,^3^ while fear of falling suggests physical and mobility-related vulnerability.^3,5^ The receipt of informal home care, conversely, points to care-related vulnerability and diminished independence.^4^ A concurrent examination of these outcomes could yield a more comprehensive perspective on potential markers associated with aging in place for Canadian seniors specifically.

### Study Objective

The present study analyzes the association between mouth-related functional problems and vulnerability among Canadian seniors. More precisely, it investigates whether discomfort during eating due to oral problems, the avoidance of foods due to mouth problems, and self-reported poor or fair oral health are associated with loneliness, fear of falling, and the receipt of informal home care. By framing oral function as a potential indicator of vulnerability, this research seeks to identify older adults at elevated risk as they age in place. The primary exposure of Experiencing Discomfort while Eating over the past 12 months (Exposure A) is expected to be associated with outcomes of high loneliness, fear of falling and receipt of informal home care.

The secondary exposures of Avoiding Foods due to mouth problems in the past 12 months (Exposure B) and Poor/Good perceived oral health (Exposure C) are also expected to be associated with fear of falling and high loneliness.

## Methods

This study utilizes cross-sectional survey data from the Canadian Health Survey on Seniors (CHSS)^19^ for secondary analysis. The CHSS survey serves as a supplement for the Canadian Community Health Survey (CCHS)^20^ and collected responses regarding the health and well-being of Canadian seniors aged 65 years or older. The survey collected data on health status, healthcare access, receipt of care, health issues, and social determinants. Data from approximately 42,000 Canadian Seniors over the 2019/2020 reference period were collected. The CHSS sampling population comprised community-dwelling adults aged 65 years and older, living in one of the ten Canadian provinces. The study excluded those who were institutionalized, members of the Canadian Armed Forces, individuals living on indigenous reserves or settlements, and those living in designated northern Quebec regions. This study used CHSS Public Use Microdata File (PUMF) data for analysis. Survey weights provided by the PUMF were used to generate weighted estimates of sample data, and CHSS-provided bootstrap weights were used for variance estimation and 95% confidence intervals.

### Measures and Variables

This study aims to evaluate mouth-related functional problems and their impact on vulnerability outcomes in seniors. All variables utilized in this study are derived from the CHSS PUMF.

The primary exposure of analysis, designated Exposure A, is the occurrence of discomfort while eating due to mouth problems over the past 12 months (PUMF variable: ‘OH3_04’). Response values of ‘Often’ and ‘Sometimes’ were assigned to the exposed group, while responses of ‘Rarely’ or ‘Never’ were assigned to the unexposed group. Exposure B is the occurrence of avoiding foods due to mouth problems in the past 12 months (‘OH3_05’). Exposure B followed the same coding rules as exposure A. Exposure C is one’s perceived oral health (‘OH3_01’), wherein a response of ‘Poor’ or ‘Fair’ is designated exposed. Exposures B and C were used for secondary analysis against vulnerability outcomes. All exposure variables were dichotomized, taking the values of ‘Exposed’ or ‘Unexposed’.

There were three vulnerability outcome variables used for this study: Loneliness, Fear of Falling, and Receipt of Informal Home Care. High Loneliness, a Fear of Falling, and the Receipt of Informal Home Care (aging in place) are all individual outcomes indicative of senior vulnerability^3,5^. The outcome variable, Loneliness, was derived from the PUMF variable of ‘LONDVSCR’, in which scores of 6 to 9 were termed ‘High Loneliness’ and scores of 3 to 5 were termed ‘Low Loneliness’. This variable is derived by the PUMF using three loneliness-related health outcome questions, Likert-scaled from 1-3, and is therefore dichotomized into ‘High Loneliness’ and ‘Low Loneliness’ groups^21^. The Fear of Falling outcome variable has dichotomized categories of ‘No Fear’ and ‘Fear of Falling’ based on the PUMF variable ‘FALDVFOF’. The final outcome variable, receipt of informal home care, has dichotomized outcomes of ‘Yes’ and ‘No’ based on the PUMF variable of ‘CR2DVFRH’, wherein individuals who received informal home care of any form in the past 12 months, including short or long-term assistance or special medical caretakers, were assigned the outcome of ‘Yes’. Hence, ‘High Loneliness’, ‘Fear of Falling’, and ‘Yes’ were the three individual outcomes representative of vulnerability.

Lastly, composite scores for exposure to mouth-related functional problems and vulnerability were created for analysis. Exposure Composite Score (ECS), defined as the sum of occurrences of mouth-related functional problem exposures (Exposures A-C), had 4 levels: ‘No Exposure’, ‘Low Exposure’, ‘Medium Exposure’, and ‘High Exposure’. The Vulnerability Composite Score (VCS), defined as the accumulation of occurrences of vulnerability outcomes (Loneliness, Fear of Falling, and Receipt of Informal Home Care), also had 4 levels. Both variables were treated as ordinal for analysis.

### Covariates

Potential covariates that could influence the study were identified within the CHSS dataset. The following covariates were included in the study: sex (‘DHH_SEX’), household size(‘DHHDGHSZ’), household income(‘INCDGHH’), highest level of education in the household(‘EHG2DVH3’), self-rated general health(‘GENVHDI’), self-rated mental health(‘GENDVMHI’), cardiovascular condition(‘CCCDGCAR’), musculoskeletal condition (‘CCCDGSKL’), respiratory condition(‘CCCDGRSP’), and smoking status(‘SMKDVSTY’).

### Sample Exclusions

All CHSS survey non-responses for any of the three exposure variables (Exposures A-C), outcome variables (Loneliness, Fear of Falling, and Receipt of Informal Home Care), and covariates were excluded from the study sample. In addition, those who responded with a preference not to answer or values indicating uncertainty were excluded. The post-exclusion unweighted sample (n = 32,756) and PUMF-provided survey weights were then used to assemble a weighted population representation of about 5.0 million community-dwelling Canadian seniors.

### Statistical Analyses

Survey weighting was performed using the R package ‘survey,’ and the CHSS full-sample survey weight (‘FWGT’) was the primary sampling weight employed to generate a population-representative sample. CHSS-provided bootstrap replicate weights were used in a replicate-weighted survey design object utilizing ‘svrepdesign()’ with ‘type = bootstrap’ in the R software ‘survey’ package. All descriptive estimates and regression models were performed using this survey design object.

A weighted descriptive table was generated to present population demographic data, and Spearman correlation analysis was performed to assess multicollinearity between covariates and exposures.

Upon analysis of covariates, the primary analysis was performed between the primary exposure (Exposure A) and the three vulnerability outcomes (Loneliness, Fear of Falling, and Receipt of Informal Home Care). Weighted prevalences of each outcome, stratified by Exposure A, were documented, along with the changes in weighted percentage frequencies for each outcome based on different exposure levels. A main logistic regression model was then generated using R version 4.6.0 for the primary analysis of Exposure A and all covariates against the three vulnerability outcome variables. A second logistic regression model of solely the outcomes’ association with Exposure A was also generated.

Lastly, secondary analysis of all exposures (Exposures A, B and C) as well as ECS, was carried out to assess effects on all three individual vulnerability outcomes and VCS. Weighted prevalences of each outcome are stratified by Exposure B, C, and ECS separately, and again assessed for changes in weighted percent frequencies. Logistic regression models (LRMs) were generated for each exposure variable, as well as combinatorial LRMs of combinations of exposures to assess effects on vulnerability outcomes. Ordinal logistic regression models (OLRs) were generated for ECS and all three exposures to assess VCS association with exposure.

## Results

### Sample Characteristics

A total of 32,756 respondents were included in this study. Characteristics of the survey-weighted population representation are shown in Table 1, grouped by exposure variables and covariates. Covariates include sex, household size, household income, smoking status, respiratory condition, musculoskeletal condition, cardiovascular condition, self-rated general health, self-rated mental health, and education. A heatmap (Figure 1) was generated depicting the Spearman correlation values for all exposure variables and covariates. Pairwise Spearman correlations among exposures and covariates were generally weak, with the strongest correlation observed between household income and household education (ρ = 0.35), suggesting no major concern for problematic collinearity.

**Table 1.**
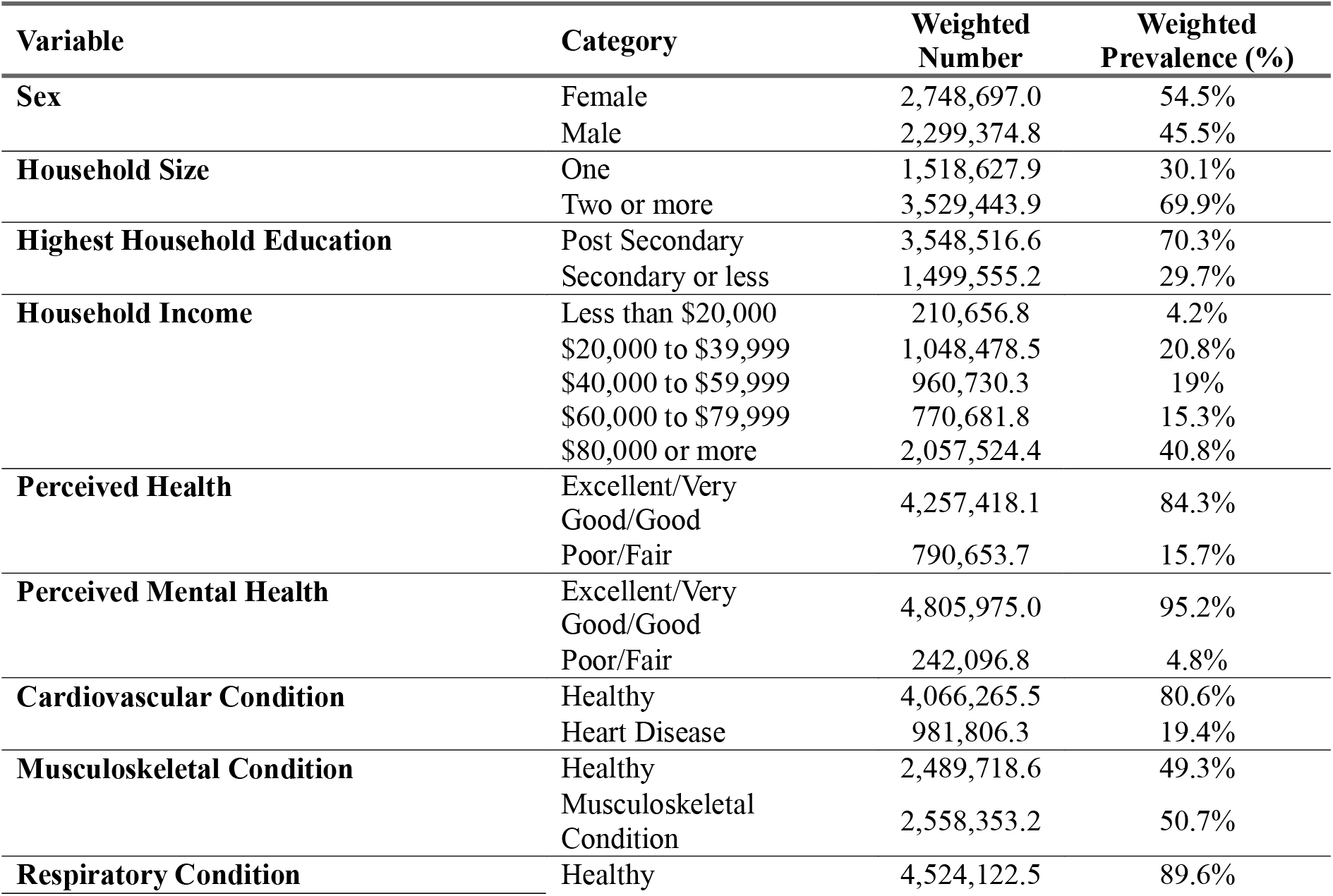
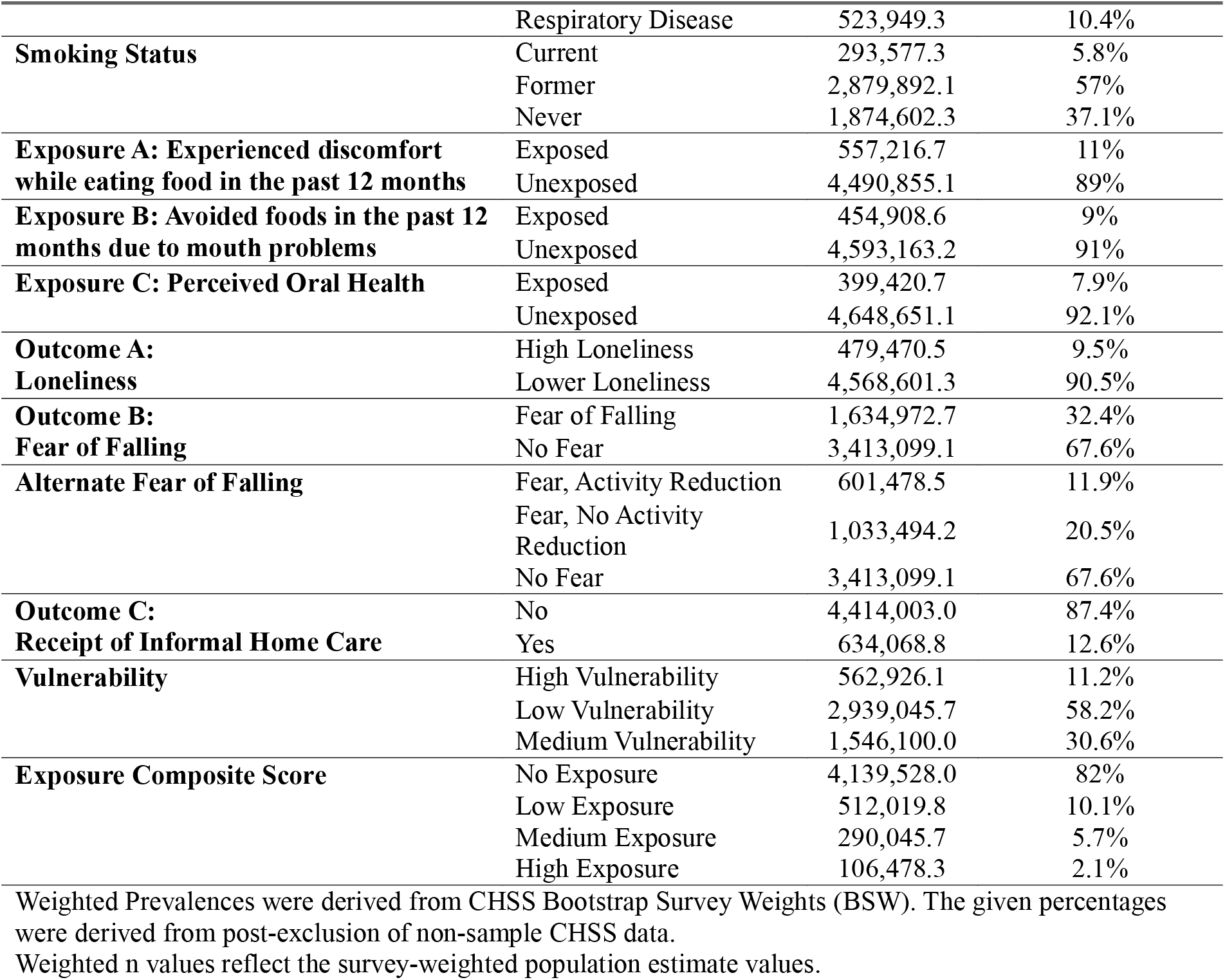
Weighted sample characteristics, CHSS.

**Figs 1-3.**
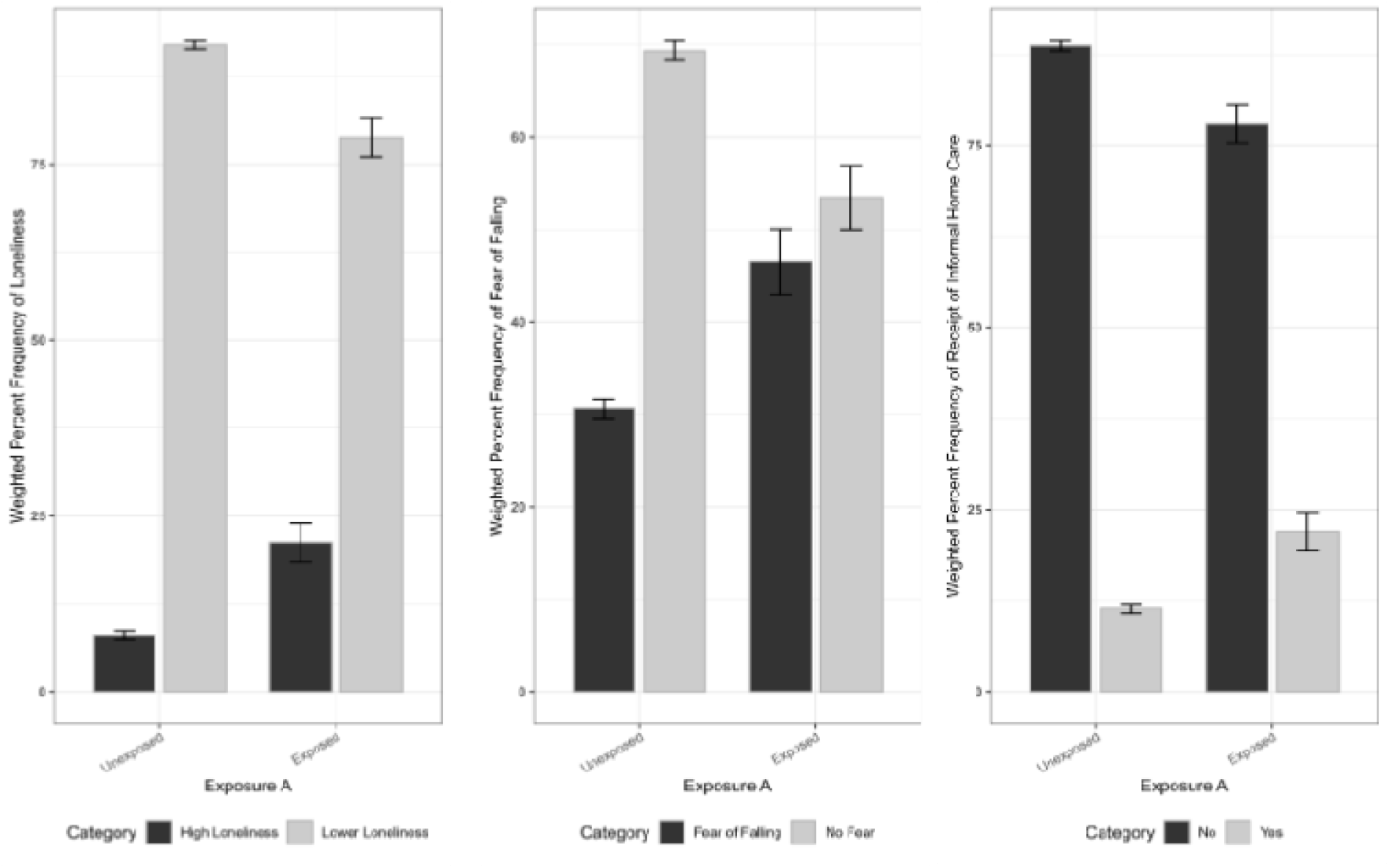
Survey-Weighted Percent Frequencies of Vulnerability Outcomes against Exposure A. The 3 panels depict the percentages of Loneliness, Fear of Falling, and Receipt of Informal Home Care among respondents who were classified as exposed or unexposed to Exposure A. Exposure A refers to those who reported experiencing discomfort while eating in the past 12 months due to mouth problems. Error bars above represent 95% Confidence Intervals.

### Primary Descriptive Findings for Exposure A

Stratified descriptive analysis of survey-weighted population data stratified by exposure variable for all variables and covariates (Exposures A-C) is presented in Tables 6, 7, and 8 (see Appendix). Table 2 presents the demographic characteristics of the sample stratified by the primary exposure (Exposure A). The outcomes specifically stratified by exposure are depicted in Table 2. The weighted prevalence of High Loneliness, Fear of Falling, and Receipt of Informal Home Care was higher among participants exposed to Exposure A than among unexposed participants (21.2% vs. 8.0%, 46.5% vs. 30.6%, and 22.1% vs. 11.4%, respectively). These results are presented visually in Figures 1-3.

**Table 2.** Weighted Percentages of Vulnerability Outcomes Stratified by Primary Exposure (Exposure A)

| Outcome | Category | Primary Exposure | Weighted % | SE | 95% CI |
| --- | --- | --- | --- | --- | --- |
| Loneliness | High Loneliness | Unexposed | 8.0 | 0.29 | 7.474-8.613 |
| Loneliness | High Loneliness | Exposed | 21.2 | 1.41 | 18.455-23.985 |
| Loneliness | Lower Loneliness | Unexposed | 92.0 | 0.29 | 91.387-92.526 |
| Loneliness | Lower Loneliness | Exposed | 78.8 | 1.41 | 76.015-81.545 |
| Fear of Falling | Fear of Falling | Unexposed | 30.6 | 0.54 | 29.576-31.692 |
| Fear of Falling | Fear of Falling | Exposed | 46.5 | 1.78 | 43.032-50.017 |
| Fear of Falling | No Fear | Unexposed | 69.4 | 0.54 | 68.308-70.424 |
| Fear of Falling | No Fear | Exposed | 53.5 | 1.78 | 49.983-56.968 |
| Receipt of Informal Home Care | No | Unexposed | 88.6 | 0.34 | 87.953-89.282 |
| Receipt of Informal Home Care | No | Exposed | 77.9 | 1.33 | 75.335-80.553 |
| Receipt of Informal Home Care | Yes | Unexposed | 11.4 | 0.34 | 10.718-12.047 |
| Receipt of Informal Home Care | Yes | Exposed | 22.1 | 1.33 | 19.447-24.665 |

Weighted Prevalences were derived from CHSS Bootstrap Survey Weights (BSW). The given percentages were derived upon exclusion of non-sample CHSS data.

### Primary Adjusted Model for Exposure A

A primary adjusted logistic regression model was derived to assess the effects of the primary exposure (Exposure A) on the three vulnerability outcomes. Model 1 (Table 3) is a logistic regression model of Exposure A, including all covariates, to assess the relationship between the outcomes and Exposure A. Odds Ratios, 95% confidence intervals, and p-values were obtained and presented in Table 3. The association with Exposure A was statistically significant (p<0.05; OR_1_ = 2.45, OR_2_ = 1.64, OR_3_ = 1.63) (Table 3).

**Table 3.** Model 1: Multivariable Logistic Regression Model of the Primary Exposure and all covariates against the Vulnerability Outcomes of Loneliness, Fear of Falling, and Receipt of Informal Home Care. Smoking status odds ratios are relative to current smokers, and household size is relative to those who live in a household of one individual.

| Characteristic | Loneliness |  |  | Fear of Falling |  |  | Receipt of Informal Home Care |  |  |
| --- | --- | --- | --- | --- | --- | --- | --- | --- | --- |
|  | OR | 95% CI | p-value | OR | 95% CI | p-value | OR | 95% CI | p-value |
| **Exposure A** | 2.45 | 2.00, 3.00 | **<0.001** | 1.64 | 1.40, 1.92 | **<0.001** | 1.63 | 1.36, 1.96 | **<0.001** |
| **Sex (male)** | 0.65 | 0.55, 0.78 | **<0.001** | 0.59 | 0.52, 0.65 | **<0.001** | 0.52 | 0.45, 0.60 | **<0.001** |
| **Household Income** |  |  |  |  |  |  |  |  |  |
| \$20,000–\$39,999 | 0.73 | 0.54, 0.99 | **0.042** | 1.07 | 0.85, 1.34 | 0.575 | 1.26 | 0.97, 1.63 | 0.081 |
| \$40,000–\$59,999 | 0.77 | 0.56, 1.06 | 0.107 | 1.02 | 0.80, 1.29 | 0.896 | 1.08 | 0.82, 1.42 | 0.600 |
| \$60,000–\$79,999 | 0.81 | 0.57, 1.15 | 0.238 | 1.03 | 0.81, 1.31 | 0.808 | 0.92 | 0.68, 1.24 | 0.582 |
| \$80,000 or more | 0.77 | 0.55, 1.09 | 0.141 | 1.15 | 0.90, 1.46 | 0.253 | 0.84 | 0.63, 1.12 | 0.247 |
| **Household Size (two or more)** | 0.28 | 0.24, 0.33 | **<0.001** | 0.74 | 0.67, 0.83 | **<0.001** | 0.59 | 0.51, 0.67 | **<0.001** |
| **Heart Disease** | 0.91 | 0.76, 1.10 | 0.345 | 1.13 | 0.99, 1.28 | 0.074 | 1.67 | 1.44, 1.93 | **<0.001** |
| **Respiratory Disease** | 1.06 | 0.87, 1.29 | 0.555 | 1.12 | 0.94, 1.32 | 0.206 | 1.34 | 1.11, 1.62 | **0.002** |
| **Musculoskeletal Disease/Condition** | 1.11 | 0.96, 1.29 | 0.165 | 1.70 | 1.54, 1.88 | **<0.001** | 1.62 | 1.41, 1.87 | **<0.001** |
| **Perceived Health (Poor/Fair)** | 1.71 | 1.40, 2.11 | **<0.001** | 1.82 | 1.57, 2.12 | **<0.001** | 2.53 | 2.14, 2.99 | **<0.001** |
| **Perceived Mental Health (Poor/Fair)** | 3.78 | 2.89, 4.95 | **<0.001** | 1.66 | 1.32, 2.10 | **<0.001** | 1.08 | 0.83, 1.42 | 0.561 |
| **Smoking Status** |  |  |  |  |  |  |  |  |  |
| Current Smoker | 1.33 | 0.95, 1.86 | 0.096 | 0.87 | 0.67, 1.12 | 0.266 | 0.68 | 0.51, 0.91 | **0.009** |
| Former Smoker | 1.04 | 0.88, 1.23 | 0.610 | 1.05 | 0.94, 1.17 | 0.380 | 0.98 | 0.85, 1.13 | 0.794 |
| **Post-Secondary Education** | 1.31 | 1.12, 1.54 | **0.001** | 1.15 | 1.03, 1.29 | **0.010** | 1.00 | 0.87, 1.14 | 0.952 |

Abbreviations: CI = Confidence Interval, OR = Odds Ratio. Bold p-values indicate statistical significance at $\alpha < 0.05$ .
Reference Categories: Female Sex, Household Income < \$20,000, Household Size of 1, No Heart/Respiratory/Musculoskeletal Disease, Excellent/Very Good/Good Perceived Health/Mental Health, Never smoked, no post-secondary education.

Model 2 (Table 4) is an unadjusted logistic model of the 3 vulnerability outcomes with the primary exposure (Exposure A) as the sole predictor. Odds Ratios, 95% confidence intervals, and p-values were obtained and presented in table 4. Consistent with the previous model, the association with Exposure A reached statistical significance (p<0.05; OR_1_ = 3.08. OR_2_ = 1.97, OR_3_ = 2.20) (Table 4).

**Table 4.** Model 2: Logistic Regression Model of Exposure A on Vulnerability Outcomes of Loneliness, Fear of Falling, and Receipt of Informal Home Care.

| Characteristic | Loneliness |  |  | Fear of Falling |  |  | Receipt of Informal Home Care |  |  |
| --- | --- | --- | --- | --- | --- | --- | --- | --- | --- |
|  | OR | 95% CI | p-value | OR | 95% CI | p-value | OR | 95% CI | p-value |
| Exposure A | 3.08 | 2.58, 3.68 | <0.001 | 1.97 | 1.69, 2.29 | <0.001 | 2.20 | 1.87, 2.60 | <0.001 |

Abbreviations: CI = Confidence Interval, OR = Odds Ratio

### Secondary Findings for Other Exposures and Composite Score

Secondary Analysis of exposure and covariates was performed by analyzing Exposure B, Exposure C, and the generated Exposure Composite Score (ECS). Descriptive models and graphical representations for Exposures B and C are provided in the appendix.

Weighted prevalence values were denoted for the three outcome variables stratified by Exposure B and Exposure C individually (Tables A1-2, Figures A2-7). Exposure to Exposure B (% frequency = 22.1, 48.1, 23.5) exhibited greater percent frequencies of the three vulnerability outcomes than no exposure (% frequency = 8.2, 30.8, 11.5). Exposure to Exposure C (% frequency = 21.0, 45.6, 21.8) again exhibited greater percent frequencies of the three vulnerability outcomes than no exposure (% frequency = 8.5, 31.2, 11.8). Descriptive analysis of the weighted population representation stratified by ECS shows a graded trend: the percentage frequencies of vulnerability outcomes increase with higher exposure scores (Tables A3-4, Figs A8-10, see supplement).

A total of 7 logistic regression models and 2 ordinal logistic regression models were created for secondary analysis and presented in tables A5-13. Model 4 (Table A6) is an ordinal regression model of all exposure variables (Exposure A, Exposure B, and Exposure C) to assess the relationship between the Vulnerability Composite Score (VCS) and each exposure. Model 5 (Table A7) utilizes ordinal logistic regression to assess associations of VCS to the Exposure Composite Score (ECS). Model 6 (Table A8) is a logistic regression model that includes all exposure variables (Exposures A-C) and covariates, and all 3 vulnerability outcome variables, to assess associations between the outcome variables and all exposures and covariates. Model 7 (Table A9) and Model 8 (Table A10) are logistic regression models of Exposure B and Exposure C, respectively, assessing the relationship between the vulnerability outcomes and each individual exposure. Models 9-11 (Tables A11-13) are logistic regression models of combinations of 2 exposure variables grouped together to assess the relationship between the three vulnerability outcome variables on each combination of exposure variables. All logistic regression models yielded odds ratios, 95% confidence intervals, and p-values. All odds ratios obtained for exposures against the three vulnerability outcomes were greater than 1.0 in all 9 secondary analysis models.

## Discussion

The study’s results indicate that for each of the three exposure measures, seniors who experienced discomfort when eating, avoided certain foods because of oral health problems, or rated their oral health as poor or fair generally demonstrated higher weighted percentages and elevated odds of reporting high loneliness, fear of falling, and reliance on informal home care. The exposure composite score further revealed a graded relationship, whereby a greater accumulation of mouth-related functional difficulties corresponded with higher levels of vulnerability across psychosocial, physical, and care-related domains. These patterns collectively imply that functional oral health problems may be indicative of wider vulnerability among community-dwelling Canadians.

### Exposure A

Exposure A, defined as experiencing discomfort while eating due to oral health issues in the preceding year, demonstrated a pronounced association with all three vulnerability measures. Both the survey-weighted descriptive analysis and the logistic regression models confirmed this relationship. According to the descriptive findings, respondents with eating discomfort had a substantially higher prevalence of loneliness, fear of falling, and informal home care. It is to be noted that these relationships persisted after adjustment. While the adjusted model yielded somewhat lower odds ratios than the unadjusted model for Exposure A, a statistically significant association with elevated odds for all three vulnerability outcomes persisted after covariate adjustment. This indicates that eating-related oral discomfort may not be merely an isolated oral health symptom but could instead signal a functional limitation linked to broader vulnerability among older adults.

### Exposure B

Exposure B, which refers to the practice of avoiding certain foods over the preceding year due to oral health issues, showed a robust association with heightened vulnerability among older adults. Those who reported avoiding foods appeared to have a stronger link with the vulnerability outcomes. In the ordinal logistic regression model constructed to predict the Vulnerability Composite Score, for instance, the odds ratio for Exposure B marginally exceeded that of Exposure A. This pattern suggests that behavioural food avoidance may represent a particularly salient functional outcome of deteriorating oral health and its subsequent vulnerability. The consistent findings from descriptive statistics, logistic regression models, and the ordinal logistic regression collectively reinforce the connection between Exposure B and elevated vulnerability. Because of this accentuated association, avoiding foods due to mouth problems could serve as a more consistent marker of senior vulnerability associated with aging in place of the three exposure variables analyzed.

### Exposure C

Exposure C, referring to respondents who reported having poor or fair self-reported oral health on the CHSS survey, exhibited an association with Loneliness and Fear of falling, while having less strength of association with receipt of informal home care. Although descriptive analysis of weighted percentages showed an increase in all three vulnerability outcomes among respondents with poor or fair self-reported oral health, the regression trend appears less robust, likely because it is a broader subjective measure.

### Exposure Composite Score

The Exposure Composite Score measured the cumulative effect of mouth-related functional difficulties across Exposures A, B, and C. Among the study’s measures, this composite indicator displayed one of the most distinct patterns. Descriptive analysis revealed a graded trend, with weighted percentages of high loneliness, fear of falling, and receipt of informal home care increasing progressively with higher exposure scores. Such a trend indicates that higher levels of vulnerability may be observed among individuals with greater accumulation of mouth-related functional difficulties. Supporting this interpretation, the ordinal logistic regression model demonstrated that a higher Exposure Composite Score was associated with increased odds of a higher Vulnerability Composite Score. Taken together, these results imply that the combined burden of multiple mouth-related functional problems may provide more meaningful information than any individual oral health measure considered in isolation.

### Associations with current literature

The associations throughout the study are similar to those in the current literature and research. The general association between experiencing discomfort while eating and avoiding food due to mouth problems in seniors aligns with the concept of oral functional decline, wherein increases in oral health problems and functional difficulties may signal a broader theme of frailty-associated vulnerability in senior populations.^22–24^

Past research has linked functional problems such as chewing difficulty, swallowing difficulty, oral hypofunction, poor oral hygiene, and poor overall oral health to impaired physical function and mortality, both of which indicate vulnerability in the senior population.^13,14^ Community-dwelling seniors have been shown to present a decline in oral function as an array of malnutritional consequences, these being greatly associated with poor overall health and vulnerability within these populations.^25–27^ Recent literature presents oral function as a potential indicator of nutrition-deficit-related vulnerability among seniors, which can manifest as general themes of social isolation or greater care needs while aging in place.^28–30^ This study finds aligned associations within the Canadian senior population, using survey-weighted CHSS data, between oral functional problems and signs of vulnerability.

Functional mobility issues, generally speaking, are a factor for those aging in place, whether due to a general loss of mobility or oral health issues caused by functional mouth problems.^5,16,18,24^ Literature related to sarcopenic dysphagia suggests that oral frailty may imply increased vulnerability and perceived vulnerability in seniors due to increased likelihood of fractures and falls.^24^ Additional research on oral health among those aging in place suggests that the progression of oral functional problems is associated with falls, thus prompting fear of falling among the senior populations. The association between fear of falling, the functional mobility indicator used in the analysis for seniors, and mouth-related functional problems (Exposure A and Exposure B) is consistent with current literature.

Additionally, associations between chewing difficulty and deteriorated oral health have been linked to an overarching theme of social isolation in past literature.^3,29,31^ Findings from previous studies suggest that deteriorating oral health or the accumulation of oral health problems may be associated with greater feelings of loneliness and isolation among adults, though stronger longitudinal evidence is required to determine the directionality of these findings.^29,31^ This study finds similar results in that mouth-related functional problems of eating discomfort and avoidance of food due to mouth problems, in addition to poorer perceived oral health, were all associated with outcomes of higher loneliness among Canadian senior populations. Overall, social isolation, a problem pertinent to Canada’s aging population, could be related to oral function problems, and this study’s findings align with previous literature in associating the two.

The increased needs associated with aging in place suggest that community-dwelling seniors with poor oral function may be receiving insufficient care to meet their general needs.^4,13,27,29^ Further caregiver research is suggestive that care while aging in the community due to greater care could be either a consequence or cause of deteriorated oral function, though directionality between the two is not currently defined.^13,18^ Additionally, a decline in oral function among those aging in place was associated with vulnerability over a two-year span, presenting a long-term association between impaired oral function and aging in place.^30^ The findings of this study suggest that exposure to mouth-related functional problems and receipt of informal home care are associated with one another, and those with mouth-related functional problems may have an increased likelihood of receiving informal home care.

Lastly, oral health research on elderly populations suggests that accumulation of oral health deficits, such as edentulism and gingivitis, may be associated with outcomes of vulnerability such as physical frailty and morbidity.^32^ Other research indicates that when oral frailty is characterized by the accumulation of oral health and functional deficits, it is associated with higher odds of nutritional vulnerability and conditions of physical frailty and sarcopenia.^23,24^ The graded trend seen within past literature of greater accumulation of oral problems being associated with higher odds of vulnerability outcomes is corroborated by the findings of this study, suggesting a graded increase in odds of vulnerability outcomes and their accumulation due to exposure to functional mouth problems.

### Strengths and Limitations

A major strength of this study is its focus on a Canadian senior population, using a large-scale sample of 32,756 respondents and approximately 5.0 million survey-weighted estimates to conduct large-scale analysis on this population. Because of this, associations found in this study are generalizable to the population of community-dwelling older Canadian adults captured by the CHSS framework. The use of CHSS-provided bootstrap weights enabled a complex study design, and the inclusion of potential or probable covariates strengthened the regression models. The use of multiple mouth-related functional problem exposures and senior vulnerability outcomes allowed for diverse analyses, along with the creation of composite scores to examine patterns associated with the accumulation of exposures and outcomes.

The primary limitation of this study is that it utilizes cross-sectional design, wherein exposure and outcome data are collected together. Consequently, the study is limited in its ability to determine causality or temporal order relationships between exposure variables and outcomes. Due to self-reported survey values and lack of confirmation by clinical examination, this is study is susceptible to recall bias, interpretation differences, or reporting biases. Due to limitations in the number of mouth-related functional problems examined, the study’s scope may be limited in the definition of mouth-related functional problems. Additionally, residual confounding may still remain despite adjustment for multiple sociodemographic and health-related covariates.

## Conclusions

The findings suggest that mouth-related functional problems, including discomfort when eating, food avoidance due to mouth problems, and poor perceived oral health, are associated with higher vulnerability among community-dwelling older Canadians. These oral functional indicators may help identify older adults at higher risk of psychosocial, physical, and support-related vulnerability relevant to aging in place.

## Data Availability

All data produced in the present study are available upon reasonable request to the authors.

