## Appendix for "Mouth-related functional problems and vulnerability among community-dwelling older Canadians: evidence from the Canadian Health Survey on Seniors"

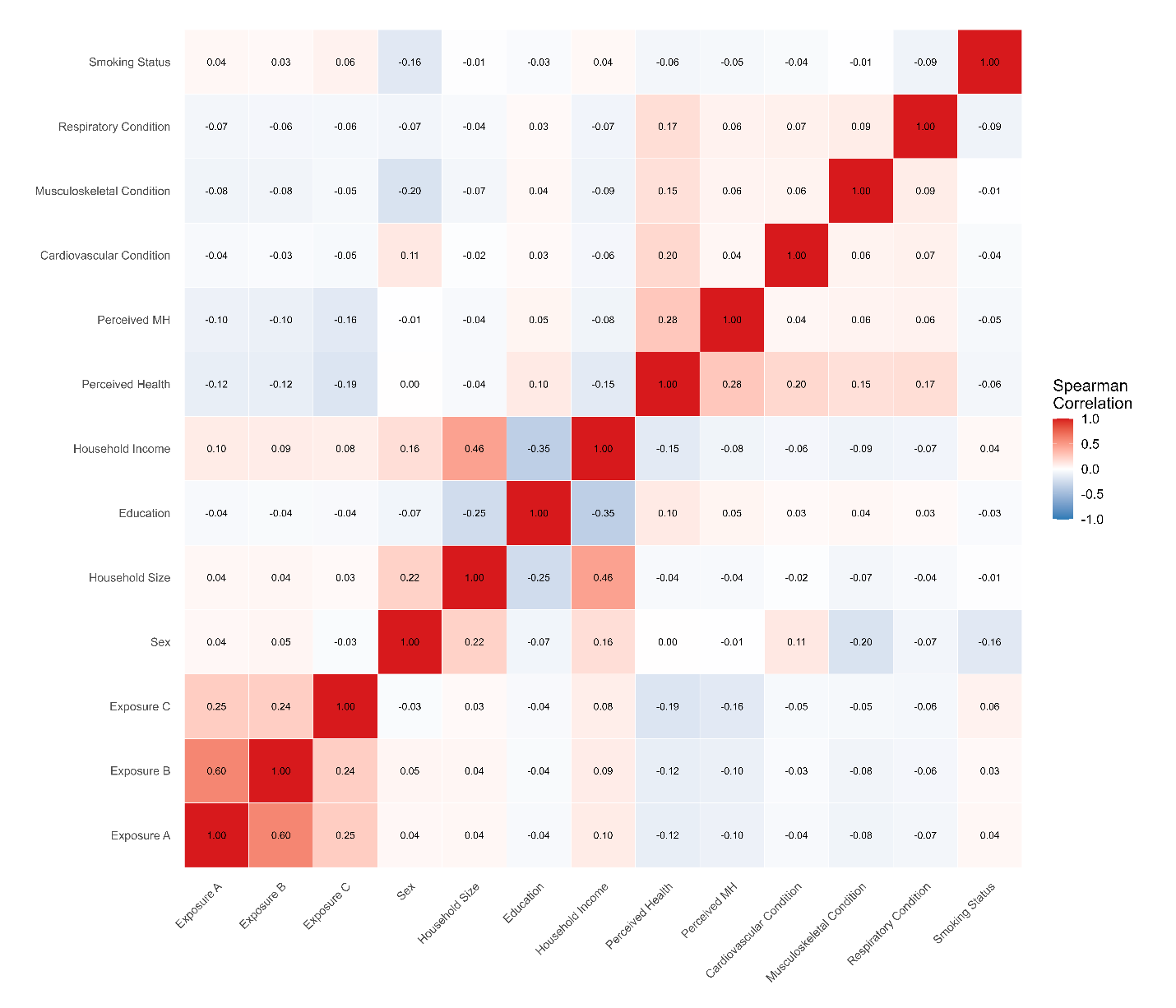

**Figure A1.** Heatmap of Spearman Correlation Values (ρ) between all Exposure Variables and Covariates to assess Multicollinearity.

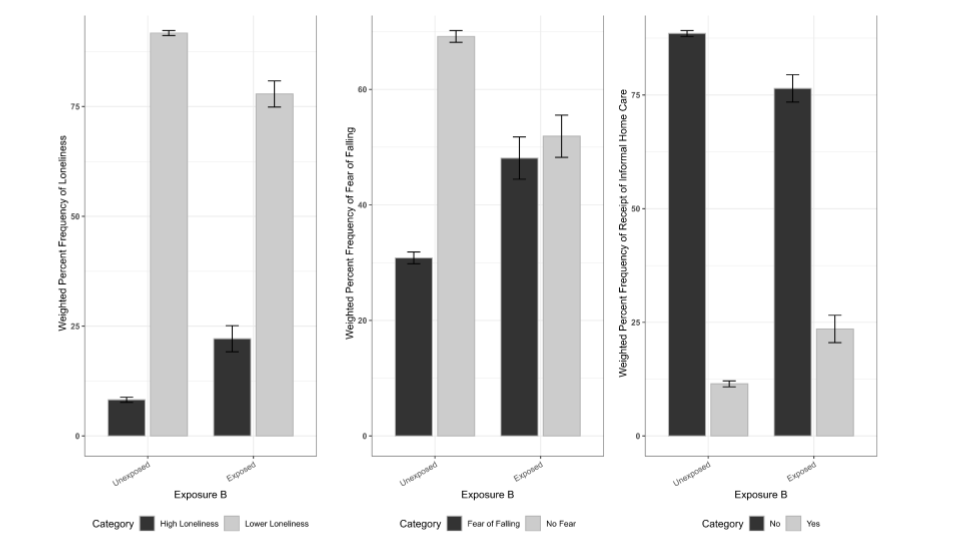

**Figs A2-4.** Survey-Weighted Percent Frequencies of Vulnerability Outcomes against Exposure B. The 3 panels depict the percent frequencies of Loneliness, Fear of Falling, and Receipt of Informal Home Care amongst respondents who were classified as exposed or unexposed to Exposure B. Exposure B refers to those who reported avoiding foods in the past 12 months due to mouth problems. Errors bars above represent 95% Confidence Intervals.

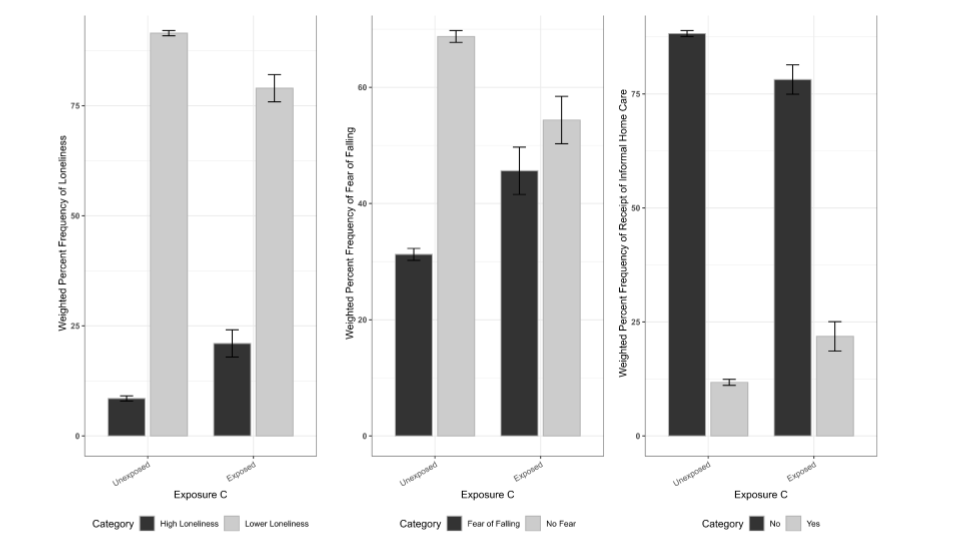

**Figs A5-7.** Survey-Weighted Percent Frequencies of Vulnerability Outcomes against Exposure C. The 3 panels depict the percent frequencies of Loneliness, Fear of Falling, and Receipt of Informal Home Care amongst respondents who were classified as exposed or unexposed to Exposure C. Exposure C refers perceived oral health as per the CHSS survey, the exposed group are those who reported having either ‘Poor’ or ‘Fair’ oral health. Errors bars above represent 95% Confidence Intervals.

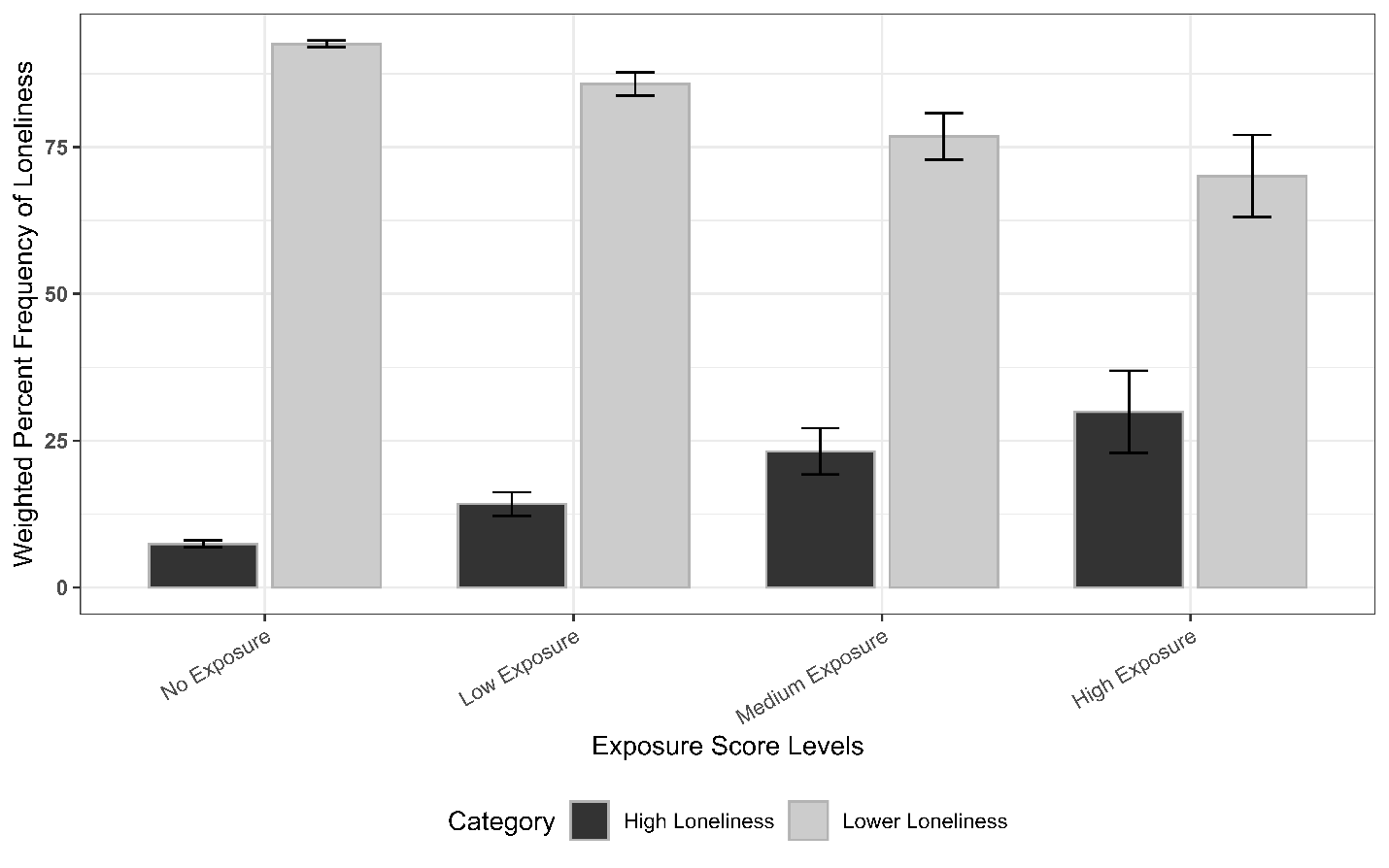

**Figure A8.** Survey-Weighted Percent Frequencies of Loneliness against Exposure Composite Score. The Exposure Composite Score has been grouped into No Exposure (score = 0), Low Exposure (score = 1), Medium Exposure (score = 2), and High Exposure (score = 3). The Exposure Score refers to mouth-related functional problems and is derived from Exposures A-C. Errors bars above represent 95% Confidence Intervals.

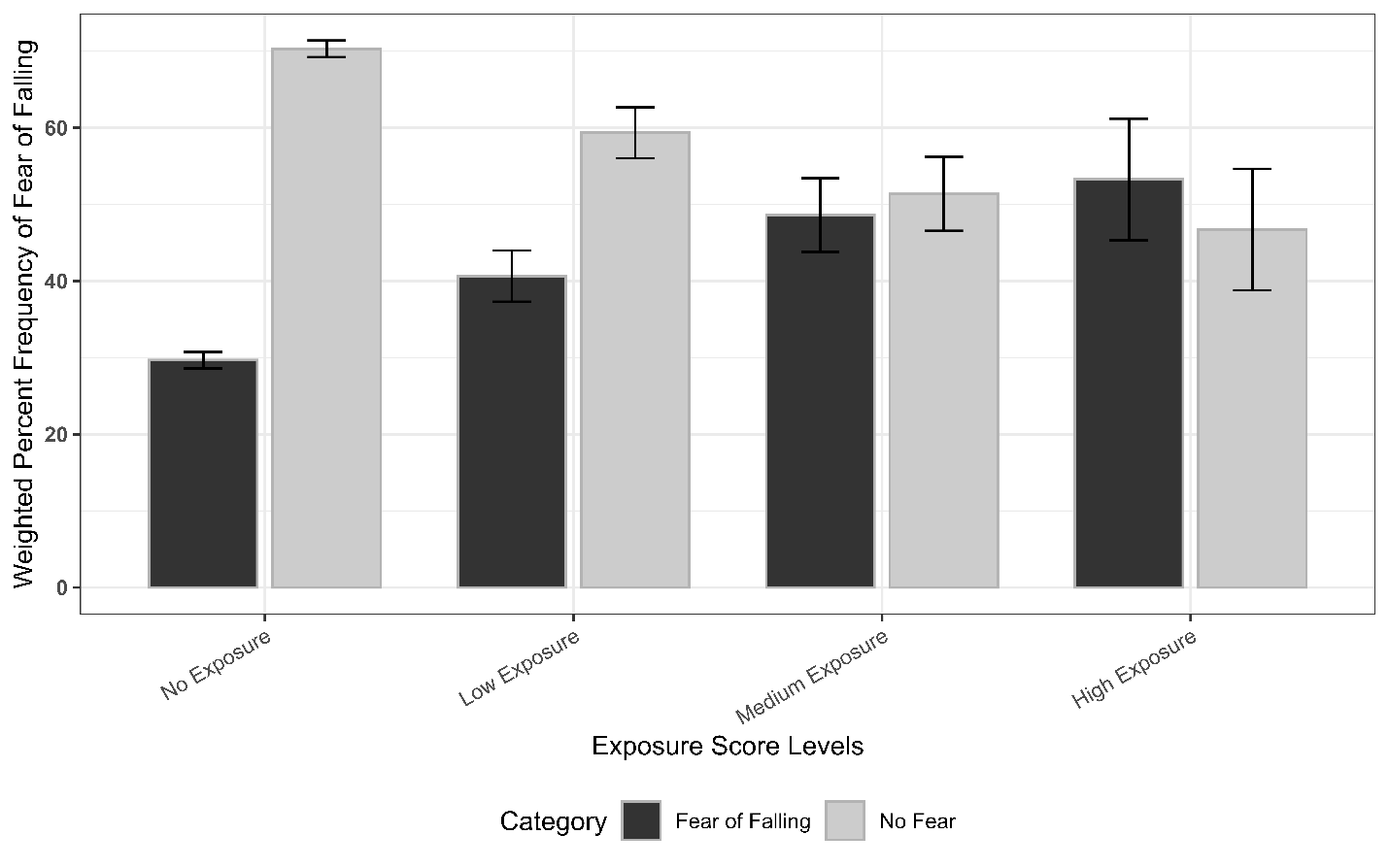

**Figure A9.** Survey-Weighted Percent Frequencies of Fear of Falling against Exposure Composite Score. The Exposure Composite Score has been grouped into No Exposure (score = 0), Low Exposure (score = 1), Medium Exposure (score = 2), and High Exposure (score = 3). The Exposure Score refers to mouth-related functional problems and is derived from Exposures A-C. Errors bars above represent 95% Confidence Intervals.

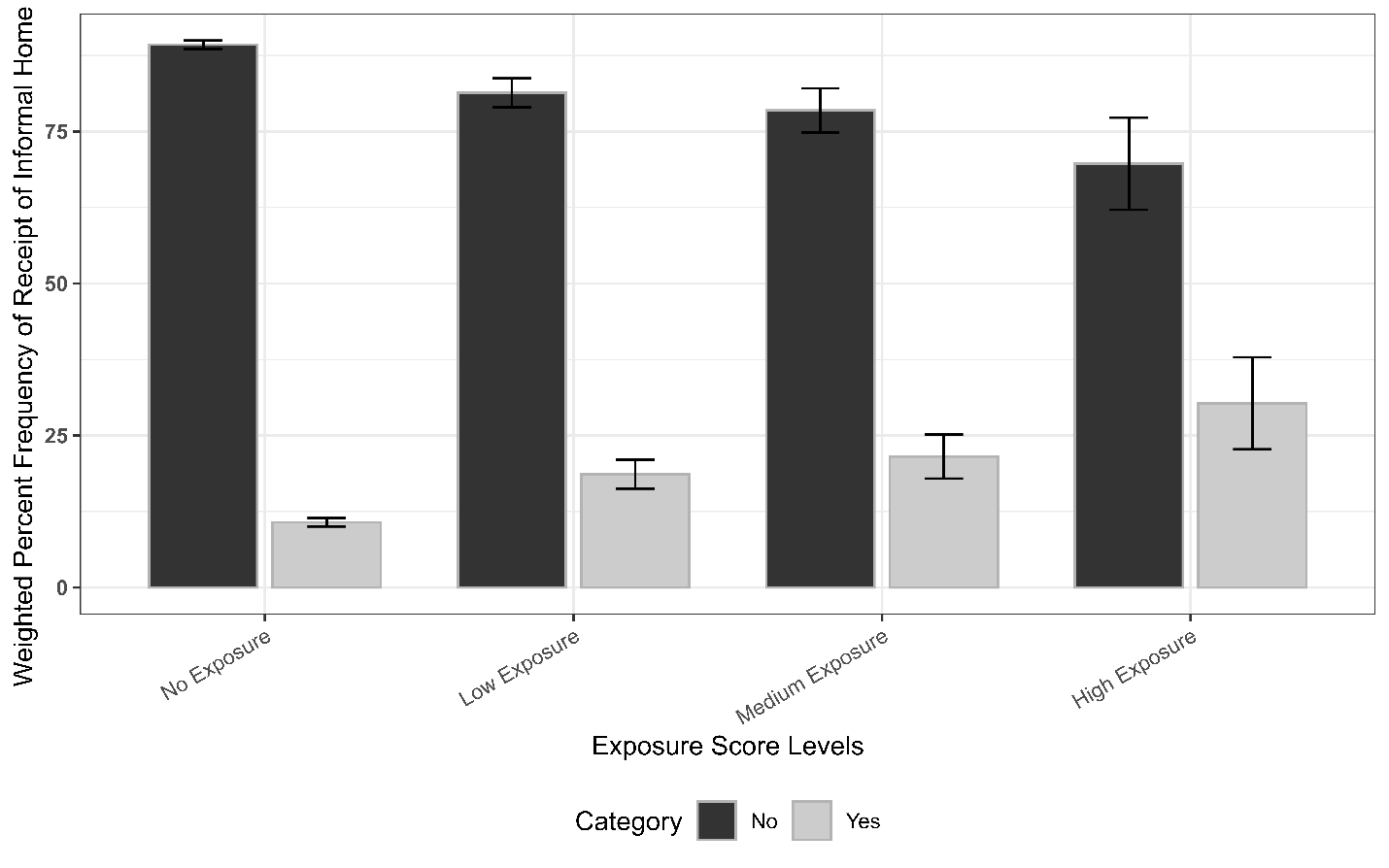

**Figure A10.** Survey-Weighted Percent Frequencies of Receipt of Informal Home Care against Exposure Composite Score. The Exposure Composite Score has been grouped into No Exposure (score = 0), Low Exposure (score = 1), Medium Exposure (score = 2), and High Exposure (score = 3). The Exposure Score refers to mouth-related functional problems and is derived from Exposures A-C. Errors bars above represent 95% Confidence Intervals.

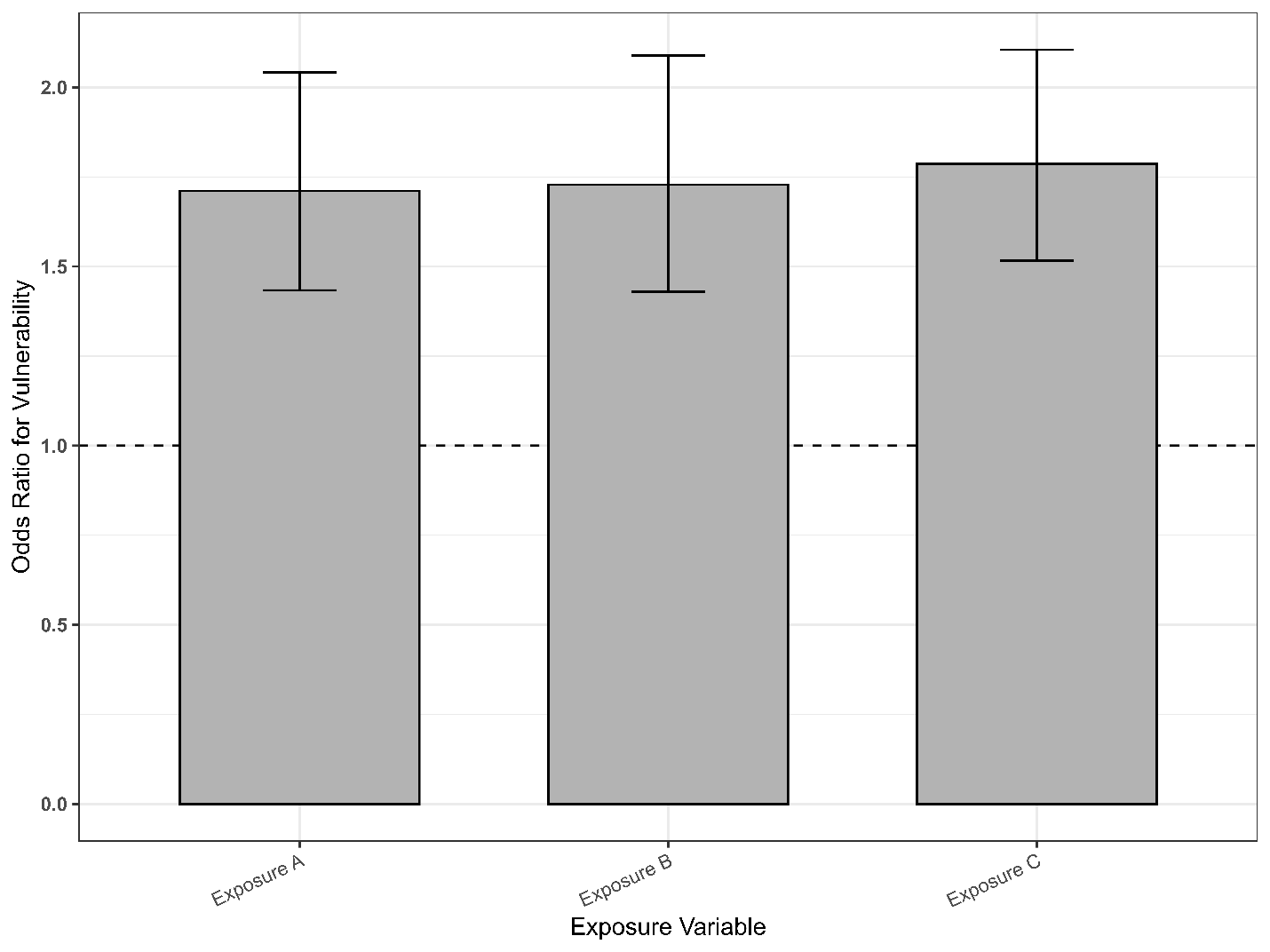

**Figure A11.** Ordinal Logistic Regression of all Exposure Variables against Vulnerability Composite Score. Errors bars represent 95% Confidence Intervals. The dashed line represents a non-correlational odds ratio threshold of 1.0.

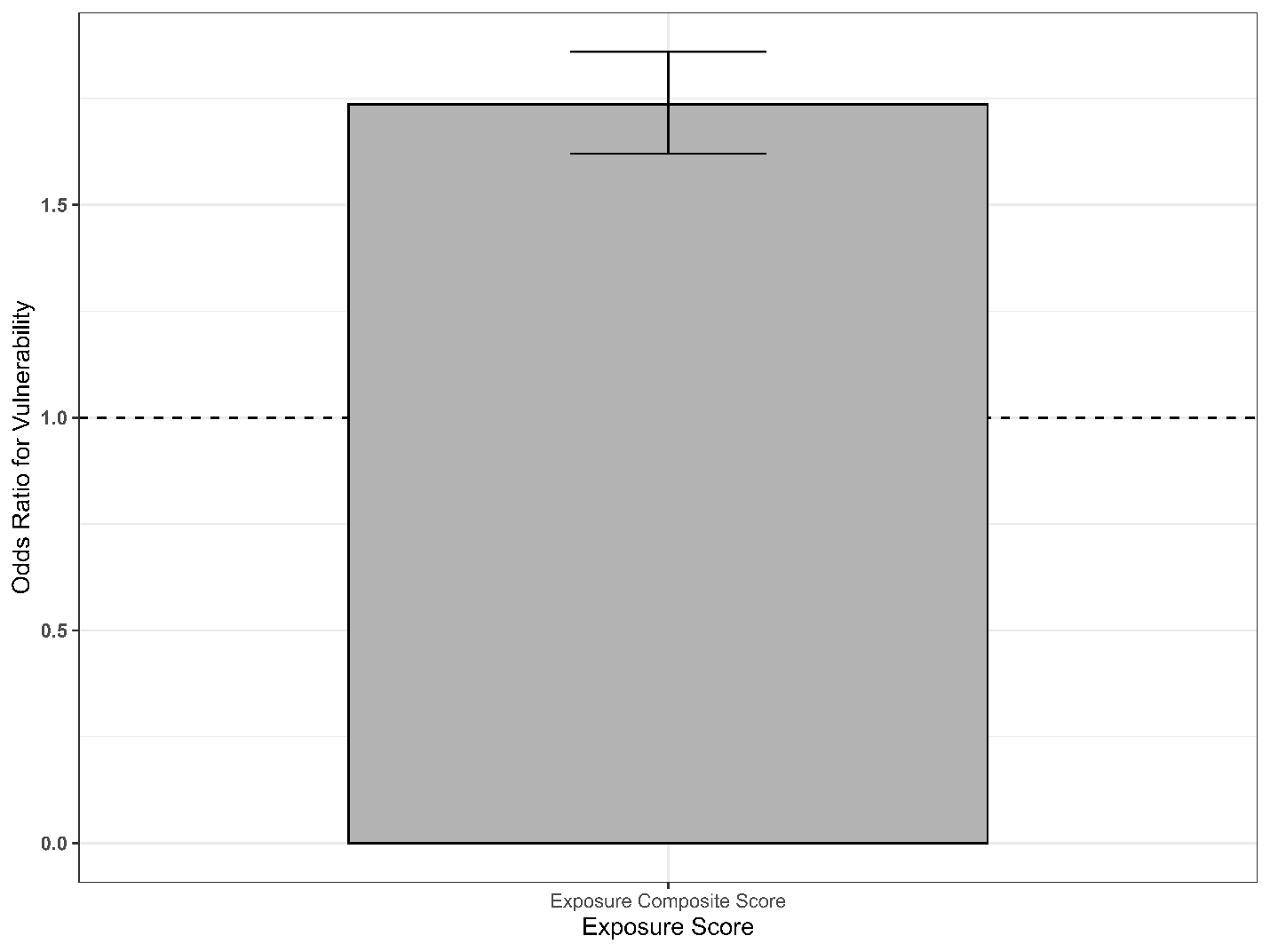

**Figure A12.** Ordinal Logistic Regression of Exposure Composite score against Vulnerability Composite Score. Error bar represents the 95% confidence interval, and the dashed line represents a non-correlational odds ratio threshold of 1.0.

Table A1. Weighted Percentages of Vulnerability Outcomes Stratified by Exposure B.

| **Outcome** | **Category** | **Exposure B** | **Weighted %** | **SE** | **95% CI** |
| --- | --- | --- | --- | --- | --- |
| Loneliness | High Loneliness | Unexposed | 8.2 | 0.29 | 7.672-8.821 |
| Loneliness | High Loneliness | Exposed | 22.1 | 1.52 | 19.159-25.112 |
| Loneliness | Lower Loneliness | Unexposed | 91.8 | 0.29 | 91.179-92.328 |
| Loneliness | Lower Loneliness | Exposed | 77.9 | 1.52 | 74.888-80.841 |
| Fear of Falling | Fear of Falling | Unexposed | 30.8 | 0.52 | 29.804-31.858 |
| Fear of Falling | Fear of Falling | Exposed | 48.1 | 1.87 | 44.449-51.764 |
| Fear of Falling | No Fear | Unexposed | 69.2 | 0.52 | 68.142-70.196 |
| Fear of Falling | No Fear | Exposed | 51.9 | 1.87 | 48.236-55.551 |
| Receipt of Informal Home Care | No | Unexposed | 88.5 | 0.33 | 87.873-89.183 |
| Receipt of Informal Home Care | No | Exposed | 76.5 | 1.54 | 73.442-79.462 |
| Receipt of Informal Home Care | Yes | Unexposed | 11.5 | 0.33 | 10.817-12.127 |
| Receipt of Informal Home Care | Yes | Exposed | 23.5 | 1.54 | 20.538-26.558 |

Table A2. Weighted Percentages of Vulnerability Outcomes Stratified by Exposure C.

| **Outcome** | **Category** | **Primary Exposure** | **Weighted %** | **SE** | **95% CI** |
| --- | --- | --- | --- | --- | --- |
| Loneliness | High Loneliness | Unexposed | 8.5 | 0.30 | 7.914-9.102 |
| Loneliness | High Loneliness | Exposed | 21.0 | 1.57 | 17.942-24.107 |
| Loneliness | Lower Loneliness | Unexposed | 91.5 | 0.30 | 90.898-92.086 |
| Loneliness | Lower Loneliness | Exposed | 79.0 | 1.57 | 75.893-82.058 |
| Fear of Falling | Fear of Falling | Unexposed | 31.2 | 0.52 | 30.223-32.277 |
| Fear of Falling | Fear of Falling | Exposed | 45.6 | 2.08 | 41.564-49.703 |
| Fear of Falling | No Fear | Unexposed | 68.8 | 0.52 | 67.723-69.777 |
| Fear of Falling | No Fear | Exposed | 54.4 | 2.08 | 50.297-58.436 |
| Receipt of Informal Home Care | No | Unexposed | 88.2 | 0.34 | 87.565-88.907 |
| Receipt of Informal Home Care | No | Exposed | 78.2 | 1.64 | 74.944-81.392 |
| Receipt of Informal Home Care | Yes | Unexposed | 11.8 | 0.34 | 11.093-12.435 |
| Receipt of Informal Home Care | Yes | Exposed | 21.8 | 1.64 | 18.608-25.056 |

Table A3. Weighted Prevalence of Covariates and Variables Stratified by Exposure Composite Score. Percentages are survey-weighted via CHSS BSWs.

| **Variable** | **Category** | **No Exposure, n(%)** | **Low Exposure, n(%)** | **Medium Exposure, n(%)** | **High Exposure, n(%)** |
| --- | --- | --- | --- | --- | --- |
| **Sex** | F | 2219269.5 (53.6%) | 283691 (55.4%) | 179131.8 (61.8%) | 66604.7 (62.6%) |
|  | M | 1920258.5 (46.4%) | 228328.8 (44.6%) | 110913.9 (38.2%) | 39873.6 (37.4%) |
| **Marital Status** | Married | 2748046.5 (66.4%) | 295096.9 (57.6%) | 168874.7 (58.2%) | 59332.3 (55.7%) |
|  | Not Married/Other | 1391481.5 (33.6%) | 216922.9 (42.4%) | 121170.9 (41.8%) | 47146 (44.3%) |
| **Household Size** | One | 1200992.5 (29%) | 174774.6 (34.1%) | 103801.6 (35.8%) | 39059.2 (36.7%) |
|  | Two or More | 2938535.5 (71%) | 337245.2 (65.9%) | 186244 (64.2%) | 67419.1 (63.3%) |
| **Highest Household Education** | Post Secondary | 2949097.7 (71.2%) | 335285 (65.5%) | 192410.2 (66.3%) | 71723.8 (67.4%) |
|  | Secondary or Less | 1190430.4 (28.8%) | 176734.8 (34.5%) | 97635.5 (33.7%) | 34754.5 (32.6%) |
| **Household Income** | $20,000 to $39,999 | 804292 (19.4%) | 135586.3 (26.5%) | 77455 (26.7%) | 31145.2 (29.3%) |
|  | $40,000 to $59,999 | 783219.7 (18.9%) | 98210.5 (19.2%) | 53616.7 (18.5%) | 25683.4 (24.1%) |
|  | $60,000 to $79,999 | 645505.6 (15.6%) | 78955.4 (15.4%) | 32462.2 (11.2%) | 13758.6 (12.9%) |
|  | $80,000 or more | 1759875.2 (42.5%) | 173210 (33.8%) | 100414.1 (34.6%) | 24025.1 (22.6%) |
|  | Less than $20,000 | 146635.5 (3.5%) | 26057.6 (5.1%) | 26097.7 (9%) | 11866 (11.1%) |
| **Perceived Health** | Excellent/Very Good/Good | 3613307.2 (87.3%) | 385242.4 (75.2%) | 206625.8 (71.2%) | 52242.7 (49.1%) |
|  | Poor/Fair | 526220.8 (12.7%) | 126777.4 (24.8%) | 83419.9 (28.8%) | 54235.6 (50.9%) |
| **Perceived Mental Health** | Excellent/Very Good/Good | 3997842.1 (96.6%) | 463883.8 (90.6%) | 262360 (90.5%) | 81889.2 (76.9%) |
|  | Poor/Fair | 141686 (3.4%) | 48136.1 (9.4%) | 27685.7 (9.5%) | 24589.1 (23.1%) |
| **Cardiovascular Condition** | Healthy | 3376527.3 (81.6%) | 392577 (76.7%) | 224327.8 (77.3%) | 72833.5 (68.4%) |
|  | Heart Disease | 763000.8 (18.4%) | 119442.8 (23.3%) | 65717.9 (22.7%) | 33644.8 (31.6%) |
| **Musculoskeletal Condition** | Healthy | 2118864 (51.2%) | 223875.7 (43.7%) | 114196.2 (39.4%) | 32782.7 (30.8%) |
|  | Musculoskeletal Condition | 2020664 (48.8%) | 288144.1 (56.3%) | 175849.4 (60.6%) | 73695.6 (69.2%) |
| **Respiratory Condition** | Healthy | 3741876 (90.4%) | 445023.6 (86.9%) | 248955 (85.8%) | 88267.9 (82.9%) |
|  | Respiratory Disease | 397652 (9.6%) | 66996.2 (13.1%) | 41090.6 (14.2%) | 18210.4 (17.1%) |
| **Smoking Status** | Current | 213969 (5.2%) | 37551.2 (7.3%) | 25437.3 (8.8%) | 16619.9 (15.6%) |
|  | Former | 2362968.6 (57.1%) | 310819.7 (60.7%) | 152971.6 (52.7%) | 53132.2 (49.9%) |
|  | Never | 1562590.4 (37.7%) | 163649 (32%) | 111636.8 (38.5%) | 36726.2 (34.5%) |
| **Exposure A** | Exposed | 0 (0%) | 190338.7 (37.2%) | 260399.8 (89.8%) | 106478.3 (100%) |
|  | Unexposed | 4139528 (100%) | 321681.1 (62.8%) | 29645.9 (10.2%) | 0 (0%) |
| **Exposure B: Avoided Foods in the past 12 months due to mouth problems** | Exposed | 0 (0%) | 104859.3 (20.5%) | 243571.1 (84%) | 106478.3 (100%) |
|  | Unexposed | 4139528 (100%) | 407160.6 (79.5%) | 46474.6 (16%) | 0 (0%) |
| **Exposure C: Perceived Oral Health** | Exposed | 0 (0%) | 216821.9 (42.3%) | 76120.5 (26.2%) | 106478.3 (100%) |
|  | Unexposed | 4139528 (100%) | 295198 (57.7%) | 213925.1 (73.8%) | 0 (0%) |
| **Outcome A: Loneliness** | High Loneliness | 307508.6 (7.4%) | 72870 (14.2%) | 67231.2 (23.2%) | 31860.7 (29.9%) |
|  | Lower Loneliness | 3832019.4 (92.6%) | 439149.8 (85.8%) | 222814.5 (76.8%) | 74617.6 (70.1%) |
| **Outcome B: Fear of Falling** | Fear of Falling | 1229131.6 (29.7%) | 208055 (40.6%) | 141061.3 (48.6%) | 56724.8 (53.3%) |
|  | No Fear | 2910396.5 (70.3%) | 303964.8 (59.4%) | 148984.4 (51.4%) | 49753.5 (46.7%) |
| **Alternate Fear of Falling** | Fear, Activity Reduction | 416160.4 (10.1%) | 84004.2 (16.4%) | 67288.1 (23.2%) | 34025.7 (32%) |
|  | Fear, No Activity Reduction | 812971.2 (19.6%) | 124050.8 (24.2%) | 73773.2 (25.4%) | 22699.1 (21.3%) |
|  | No Fear | 2910396.5 (70.3%) | 303964.8 (59.4%) | 148984.4 (51.4%) | 49753.5 (46.7%) |
| **Outcome C: Receipt of Informal Home Care** | No | 3695663.4 (89.3%) | 416570.2 (81.4%) | 227557.2 (78.5%) | 74212.2 (69.7%) |
|  | Yes | 443864.7 (10.7%) | 95449.6 (18.6%) | 62488.4 (21.5%) | 32266 (30.3%) |
| **Vulnerability** | High Vulnerability | 362220 (8.8%) | 87707.9 (17.1%) | 75497 (26%) | 37501.1 (35.2%) |
|  | Low Vulnerability | 2561473.9 (61.9%) | 237863.7 (46.5%) | 105612.2 (36.4%) | 34095.9 (32%) |
|  | Medium Vulnerability | 1215834.1 (29.4%) | 186448.1 (36.4%) | 108936.5 (37.6%) | 34881.2 (32.8%) |
| **Exposure Comp**  **Score** | 0 | 4139528 (100%) | 0 (0%) | 0 (0%) | 0 (0%) |
|  | 1 | 0 (0%) | 512019.8 (100%) | 0 (0%) | 0 (0%) |
|  | 2 | 0 (0%) | 0 (0%) | 290045.7 (100%) | 0 (0%) |
|  | 3 | 0 (0%) | 0 (0%) | 0 (0%) | 106478.3 (100%) |
| Exposure Composite score is the sum of all Exposures wherein ‘Exposed’ is 1 and ‘Unexposed’ is 0. | | | | | |

Table A4. Weighted Percent Frequencies of Vulnerability Outcomes Stratified by Exposure Composite Score (ECS).

| **Outcome** | **Category** | **Exposure Score Level** | **Weighted %** | **SE** | **95% CI** |
| --- | --- | --- | --- | --- | --- |
| Loneliness | High Loneliness | No Exposure | 7.4 | 0.29 | 6.85-8.007 |
| Loneliness | High Loneliness | Low Exposure | 14.2 | 1.03 | 12.222-16.242 |
| Loneliness | High Loneliness | Medium Exposure | 23.2 | 2.01 | 19.236-27.123 |
| Loneliness | High Loneliness | High Exposure | 29.9 | 3.57 | 22.928-36.916 |
| Loneliness | Lower Loneliness | No Exposure | 92.6 | 0.29 | 91.993-93.15 |
| Loneliness | Lower Loneliness | Low Exposure | 85.8 | 1.03 | 83.758-87.778 |
| Loneliness | Lower Loneliness | Medium Exposure | 76.8 | 2.01 | 72.877-80.764 |
| Loneliness | Lower Loneliness | High Exposure | 70.1 | 3.57 | 63.084-77.072 |
| Fear of Falling | Fear of Falling | No Exposure | 29.7 | 0.55 | 28.61-30.775 |
| Fear of Falling | Fear of Falling | Low Exposure | 40.6 | 1.70 | 37.293-43.975 |
| Fear of Falling | Fear of Falling | Medium Exposure | 48.6 | 2.46 | 43.815-53.454 |
| Fear of Falling | Fear of Falling | High Exposure | 53.3 | 4.04 | 45.349-61.198 |
| Fear of Falling | No Fear | No Exposure | 70.3 | 0.55 | 69.225-71.39 |
| Fear of Falling | No Fear | Low Exposure | 59.4 | 1.70 | 56.025-62.707 |
| Fear of Falling | No Fear | Medium Exposure | 51.4 | 2.46 | 46.546-56.185 |
| Fear of Falling | No Fear | High Exposure | 46.7 | 4.04 | 38.802-54.651 |
| Receipt of Informal Home Care | No | No Exposure | 89.3 | 0.35 | 88.6-89.955 |
| Receipt of Informal Home Care | No | Low Exposure | 81.4 | 1.21 | 78.986-83.73 |
| Receipt of Informal Home Care | No | Medium Exposure | 78.5 | 1.85 | 74.835-82.077 |
| Receipt of Informal Home Care | No | High Exposure | 69.7 | 3.87 | 62.118-77.276 |
| Receipt of Informal Home Care | Yes | No Exposure | 10.7 | 0.35 | 10.045-11.4 |
| Receipt of Informal Home Care | Yes | Low Exposure | 18.6 | 1.21 | 16.27-21.014 |
| Receipt of Informal Home Care | Yes | Medium Exposure | 21.5 | 1.85 | 17.923-25.165 |
| Receipt of Informal Home Care | Yes | High Exposure | 30.3 | 3.87 | 22.724-37.882 |

|  | **Loneliness** | | | **Fear of Falling** | | | **Receipt of Informal Home Care** | | |
| --- | --- | --- | --- | --- | --- | --- | --- | --- | --- |
| **Characteristic** | **OR** | **95% CI** | **p-value** | **OR** | **95% CI** | **p-value** | **OR** | **95% CI** | **p-value** |
| Exposure A | 1.92 | 1.50, 2.45 | <0.001 | 1.46 | 1.20, 1.77 | <0.001 | 1.50 | 1.21, 1.87 | <0.001 |
| Exposure B | 1.70 | 1.31, 2.21 | <0.001 | 1.48 | 1.21, 1.80 | <0.001 | 1.61 | 1.28, 2.02 | <0.001 |
| Exposure C | 1.97 | 1.58, 2.46 | <0.001 | 1.49 | 1.25, 1.77 | <0.001 | 1.60 | 1.29, 1.98 | <0.001 |
| Abbreviations: CI = Confidence Interval, OR = Odds Ratio | | | | | | | | | |

**Table A5.** Model 3: Logistic regression Model of all Exposure Variables on Vulnerability Outcomes.

| Exposure | OR | (95% CI) | SE | p-value |
| --- | --- | --- | --- | --- |
| Exposure Composite Score | 1.736 | (1.62-1.86) | 0.035 | <0.001 |

**Table A6.** Model 5: Ordinal logistic regression model of Vulnerability Composite Score Against Exposure Composite Score.

| Exposure | OR | (95% CI) | SE | p-value |
| --- | --- | --- | --- | --- |
| Exposure A | 1.711 | (1.434-2.041) | 0.090 | <0.001 |
| Exposure B | 1.728 | (1.43-2.089) | 0.097 | <0.001 |
| Exposure C | 1.786 | (1.516-2.105) | 0.084 | <0.001 |

**Table A7.** Model 4: Ordinal logistic regression of Vulnerability Composite Score against all Exposure Variables.

|  | **Loneliness** | | | **Fear of Falling** | | | **Receipt of Informal Home Care** | | |
| --- | --- | --- | --- | --- | --- | --- | --- | --- | --- |
| **Characteristic** | **OR** | **95% CI** | **p-value** | **OR** | **95% CI** | **p-value** | **OR** | **95% CI** | **p-value** |
| Exposure A | 1.80 | 1.38, 2.35 | <0.001 | 1.34 | 1.10, 1.64 | 0.004 | 1.29 | 1.04, 1.62 | 0.024 |
| Exposure B | 1.48 | 1.12, 1.97 | 0.007 | 1.34 | 1.09, 1.64 | 0.005 | 1.39 | 1.09, 1.77 | 0.007 |
| Exposure C | 1.48 | 1.15, 1.92 | 0.003 | 1.25 | 1.04, 1.49 | 0.016 | 1.22 | 0.96, 1.56 | 0.109 |
| Sex |  |  |  |  |  |  |  |  |  |
| F | — | — |  | — | — |  | — | — |  |
| M | 0.65 | 0.55, 0.78 | <0.001 | 0.59 | 0.52, 0.65 | <0.001 | 0.52 | 0.45, 0.61 | <0.001 |
| Household Income |  |  |  |  |  |  |  |  |  |
| $20,000 to $39,999 | — | — |  | — | — |  | — | — |  |
| $40,000 to $59,999 | 1.05 | 0.86, 1.27 | 0.657 | 0.95 | 0.82, 1.10 | 0.500 | 0.85 | 0.71, 1.02 | 0.080 |
| $60,000 to $79,999 | 1.13 | 0.90, 1.42 | 0.307 | 0.97 | 0.82, 1.15 | 0.754 | 0.74 | 0.60, 0.91 | 0.004 |
| $80,000 or more | 1.07 | 0.86, 1.35 | 0.529 | 1.09 | 0.93, 1.26 | 0.290 | 0.67 | 0.55, 0.83 | <0.001 |
| Less than $20,000 | 1.33 | 0.98, 1.81 | 0.068 | 0.92 | 0.73, 1.16 | 0.484 | 0.78 | 0.60, 1.01 | 0.062 |
| Household Size |  |  |  |  |  |  |  |  |  |
| One | — | — |  | — | — |  | — | — |  |
| Two or More | 0.28 | 0.24, 0.33 | <0.001 | 0.74 | 0.67, 0.83 | <0.001 | 0.58 | 0.51, 0.67 | <0.001 |
| Cardiovascular Condition |  |  |  |  |  |  |  |  |  |
| Healthy | — | — |  | — | — |  | — | — |  |
| Heart Disease | 0.91 | 0.75, 1.10 | 0.314 | 1.12 | 0.99, 1.28 | 0.077 | 1.66 | 1.43, 1.93 | <0.001 |
| Respiratory Condition |  |  |  |  |  |  |  |  |  |
| Healthy | — | — |  | — | — |  | — | — |  |
| Respiratory Disease | 1.05 | 0.86, 1.28 | 0.618 | 1.11 | 0.94, 1.32 | 0.219 | 1.34 | 1.11, 1.62 | 0.002 |
| Musculoskeletal Condition |  |  |  |  |  |  |  |  |  |
| Healthy | — | — |  | — | — |  | — | — |  |
| Musculoskeletal Condition | 1.10 | 0.95, 1.28 | 0.191 | 1.70 | 1.54, 1.88 | <0.001 | 1.62 | 1.41, 1.86 | <0.001 |
| Perceived Health |  |  |  |  |  |  |  |  |  |
| Excellent/Very Good/Good | — | — |  | — | — |  | — | — |  |
| Poor/Fair | 1.62 | 1.33, 1.98 | <0.001 | 1.77 | 1.52, 2.06 | <0.001 | 2.45 | 2.08, 2.89 | <0.001 |
| Perceived Mental Health |  |  |  |  |  |  |  |  |  |
| Excellent/Very Good/Good | — | — |  | — | — |  | — | — |  |
| Poor/Fair | 3.55 | 2.68, 4.70 | <0.001 | 1.60 | 1.27, 2.03 | <0.001 | 1.03 | 0.79, 1.36 | 0.808 |
| Smoking Status |  |  |  |  |  |  |  |  |  |
| Current | — | — |  | — | — |  | — | — |  |
| Former | 0.81 | 0.58, 1.13 | 0.207 | 1.23 | 0.95, 1.58 | 0.113 | 1.47 | 1.10, 1.96 | 0.010 |
| Never | 0.77 | 0.54, 1.08 | 0.133 | 1.17 | 0.90, 1.51 | 0.232 | 1.49 | 1.11, 2.01 | 0.008 |
| Education |  |  |  |  |  |  |  |  |  |
| Post Secondary | — | — |  | — | — |  | — | — |  |
| Secondary or Less | 0.77 | 0.66, 0.91 | 0.002 | 0.87 | 0.78, 0.97 | 0.012 | 1.01 | 0.89, 1.15 | 0.877 |
| Abbreviations: CI = Confidence Interval, OR = Odds Ratio | | | | | | | | | |

**Table A8.** Model 6**:** Logistic regression model of all Mouth-related Functional Problem Exposure Variables and Covariates against 3 Vulnerability Outcomes of Loneliness, Fear of Falling, and Receipt of Informal Home Care

|  | **Loneliness** | | | **Fear of Falling** | | | **Receipt of Informal Home Care** | | |
| --- | --- | --- | --- | --- | --- | --- | --- | --- | --- |
| **Characteristic** | **OR** | **95% CI** | **p-value** | **OR** | **95% CI** | **p-value** | **OR** | **95% CI** | **p-value** |
| Exposure B | 3.16 | 2.63, 3.80 | <0.001 | 2.08 | 1.78, 2.42 | <0.001 | 2.38 | 1.99, 2.84 | <0.001 |
| Abbreviations: CI = Confidence Interval, OR = Odds Ratio | | | | | | | | | |

**Table A9.** Model 7: Logistic regression model of Exposure B on Vulnerability Outcomes.

|  | **Loneliness** | | | **Fear of Falling** | | | **Receipt of Informal Home Care** | | |
| --- | --- | --- | --- | --- | --- | --- | --- | --- | --- |
| **Characteristic** | **OR** | **95% CI** | **p-value** | **OR** | **95% CI** | **p-value** | **OR** | **95% CI** | **p-value** |
| Exposure C | 2.86 | 2.35, 3.49 | <0.001 | 1.85 | 1.56, 2.19 | <0.001 | 2.09 | 1.71, 2.56 | <0.001 |
| Abbreviations: CI = Confidence Interval, OR = Odds Ratio | | | | | | | | | |

**Table A10.** Model 8: Logistic regression model of Exposure C on Vulnerability Outcomes.

|  | **Loneliness** | | | **Fear of Falling** | | | **Receipt of Informal Home Care** | | |
| --- | --- | --- | --- | --- | --- | --- | --- | --- | --- |
| **Characteristic** | **OR** | **95% CI** | **p-value** | **OR** | **95% CI** | **p-value** | **OR** | **95% CI** | **p-value** |
| Exposure B | 2.58 | 2.11, 3.16 | <0.001 | 1.87 | 1.60, 2.18 | <0.001 | 2.08 | 1.73, 2.49 | <0.001 |
| Exposure C | 2.14 | 1.72, 2.67 | <0.001 | 1.56 | 1.31, 1.86 | <0.001 | 1.68 | 1.37, 2.07 | <0.001 |
| Abbreviations: CI = Confidence Interval, OR = Odds Ratio | | | | | | | | | |

**Table A11.** Model 9. Logistic regression model of Exposure B and Exposure C on Vulnerability Outcomes.

|  | **Loneliness** | | | **Fear of Falling** | | | **Receipt of Informal Home Care** | | |
| --- | --- | --- | --- | --- | --- | --- | --- | --- | --- |
| **Characteristic** | **OR** | **95% CI** | **p-value** | **OR** | **95% CI** | **p-value** | **OR** | **95% CI** | **p-value** |
| Exposure A | 2.12 | 1.66, 2.71 | <0.001 | 1.54 | 1.27, 1.87 | <0.001 | 1.61 | 1.30, 1.98 | <0.001 |
| Exposure B | 1.90 | 1.47, 2.45 | <0.001 | 1.57 | 1.28, 1.92 | <0.001 | 1.73 | 1.38, 2.17 | <0.001 |
| Abbreviations: CI = Confidence Interval, OR = Odds Ratio | | | | | | | | | |

**Table A12.** Model 10. Logistic regression model of Exposure A and Exposure B on Vulnerability Outcomes.

|  | **Loneliness** | | | **Fear of Falling** | | | **Receipt of Informal Home Care** | | |
| --- | --- | --- | --- | --- | --- | --- | --- | --- | --- |
| **Characteristic** | **OR** | **95% CI** | **p-value** | **OR** | **95% CI** | **p-value** | **OR** | **95% CI** | **p-value** |
| Exposure A | 2.57 | 2.12, 3.10 | <0.001 | 1.79 | 1.54, 2.08 | <0.001 | 1.94 | 1.63, 2.31 | <0.001 |
| Exposure C | 2.11 | 1.70, 2.61 | <0.001 | 1.56 | 1.31, 1.85 | <0.001 | 1.70 | 1.37, 2.10 | <0.001 |
| Abbreviations: CI = Confidence Interval, OR = Odds Ratio | | | | | | | | | |

**Table A13.** Model 11: Logistic regression model of Exposure A and Exposure C on Vulnerability Outcomes.
